# Comparative immunogenicity of several inactivated influenza vaccines among healthcare workers in Hong Kong

**DOI:** 10.64898/2026.08.19.26360857

**Authors:** Wey Wen Lim, Lisa Touyon, Loretta Mak, Samuel M. S. Cheng, Yiu Chung Lau, Dennis K. M. Ip, Malik Peiris, Benjamin J. Cowling, Sook-San Wong

## Abstract

We compared the immunogenicity of three licensed egg-based inactivated influenza vaccines, including TetrAnflu (Sinovac quadrivalent), Fluarix Tetra (GSK quadrivalent), and Vaxigrip (Sanofi trivalent), in adult healthcare workers in Hong Kong during the 2025/26 season. Age- and sex-matched recipients (n=30–40 per group) with paired pre- and post-vaccination sera were identified from an ongoing cohort. Hemagglutination-inhibition assays were performed against the egg-propagated 2025/26 vaccine reference strains. The three vaccines induced broadly comparable geometric mean fold-rises, post-vaccination geometric mean titers, proportions achieving titers ≥40, and seroconversion rates against vaccine strains. These real-world data support the interchangeability of the three products for seasonal influenza vaccination programs among healthcare workers.

**Brief summary:** We compared the immunogenicity of three licensed egg-based inactivated influenza vaccines among age- and sex-matched healthcare workers in Hong Kong. Antibody responses were broadly comparable, supporting their interchangeability for influenza vaccination programs.

## BACKGROUND

Several vaccine manufacturers have licensed egg-based inactivated influenza vaccines [1]. In Hong Kong, egg-grown inactivated influenza vaccines produced by Sanofi, GlaxoSmithKline, Abbott, and Sinovac Biotech are licensed for use. Each year the majority of vaccines are provided through the public sector via a tender process to select one or more products, with vaccinations also available through the private sector. As in many other locations, healthcare workers in Hong Kong are recommended to receive influenza vaccination annually, and the uptake locally is around 40% to 50% [2,3]. TetrAnflu (Sinovac Biotech) quadrivalent split-virion inactivated influenza vaccine reported phase III results in 2020 and 2024 [4–6], and received approval for use in Hong Kong in May 2025. In this study we aimed to compare the immunogenicity of available inactivated influenza vaccines in young adult healthcare workers.

## METHODS

### Study participants

Between June 2020 and February 2026, we enrolled and followed up healthcare workers in Hong Kong in a longitudinal cohort study [7]. Participants provided blood samples at enrolment and then every six months, and provided additional samples at one month after receipt of any influenza vaccination or COVID-19 vaccination. Participants provided demographic and occupational information in the twice-annual follow-up visits, and reported any acute respiratory illnesses.

We selected pairs of pre-vaccination (baseline) and post-vaccination sera from participants who received influenza vaccination for the 2025/26 northern hemisphere season with either TetrAnflu (quadrivalent split-virion inactivated influenza vaccine manufactured by Sinovac Biotech), Vaxigrip (trivalent split-virion inactivated influenza vaccine manufactured by Sanofi), or Fluarix (quadrivalent split-virion inactivated influenza vaccine manufactured by GSK). The northern hemisphere vaccines included 15µg of the hemagglutinin (HA) influenza surface protein of each of the following three strains: A/Victoria/4897/2022-like virus (H1N1), A/Croatia/10136RV/2023-like virus (H3N2) and B/Austria/1359417/2021-like virus (Victoria lineage). Fluarix Tetra and TetrAnflu were quadrivalent vaccines and additionally included 15µg of the HA of a B/Phuket/3073/2013-like virus (Yamagata lineage). Pre-vaccination sera were defined as serum samples collected within 180 days before any influenza vaccination, and post-vaccination sera were defined as serum samples collected 21 to 42 days after any influenza vaccination.

Among the 906 active participants on 1 September 2025 there were 438 participants who reported 2025/26 influenza vaccination. We selected age- and sex-matched recipients of TetrAnflu (n=40), Fluarix Tetra (n=30) and Vaxigrip (n=40) who had both a pre-vaccination serum collected within 180 days before vaccination and a post-vaccination serum collected 21-42 days afterward. Matching was performed to balance age and sex across the three available products. HAI testing capacity limited the number of pairs that could be assayed.

### Laboratory methods

Paired sera were tested in parallel using hemagglutination inhibition (HAI) assays against the egg-propagated vaccine reference viruses matching the 2025/26 Northern Hemisphere formulation. Sera were tested in serial doubling dilutions, and the HAI antibody titer was taken as the reciprocal of the last dilution that showed complete inhibition of hemagglutination. HAI antibody titers of <10 were imputed as 5 for analysis.

### Statistical analysis

We described demographics, occupational information, and health statuses of participants who received the different vaccines with descriptive statistics, and compared these characteristics between groups using ANOVA tests for continuous variables and chi-square or Fisher’s tests for categorical variables. The primary comparison was the geometric mean fold-rise in titer from pre-to post-vaccination. We calculated the proportion of HAI antibody titer against each vaccine strain for all participants and the proportion of participants that have achieved an HAI antibody titer ≥40 after vaccination. We also calculated the proportion of participants who achieved seroconversion, defined as either (1) achieving a post-vaccination titer ≥40 from a pre-vaccination titer of <10; or (2) achieving a four-fold or greater rise in post-vaccination titer from a pre-vaccination titer of ≥10. We also calculated the proportion of participants that achieved an HAI antibody titer ≥40 after vaccination. We then compared the differences in mean-fold rise, the seroconversion rate, and the proportion of participants achieving a post-vaccination titer ≥40 between the three vaccine groups. We conducted crude comparisons, given that groups were already matched in the selection stage, and we additionally performed analyses adjusting for age, sex, and days from pre-vaccination sampling to vaccination. We performed a sensitivity analysis restricted to participants whose pre-sample was obtained within 90 days of vaccination.

### Ethical approval

The study received ethical approval from the Institutional Review Board of the University of Hong Kong/Hospital Authority Hong Kong West Cluster Institutional Review Board (ref: UW 20-344). All participants provided written informed consent to participate.

## RESULTS

Between 29 June 2020 and 22 October 2024, we recruited 1,758 healthcare workers from healthcare institutions across Hong Kong and followed them up to 28 February 2026. There were 906 active participants in the cohort on 1 September 2025, and 438 (48.3%) reported receipt of an influenza vaccination between September and December 2025. Of these, 222 (50.6%) reported receipt of Vaxigrip trivalent (Sanofi), 94 (42.3%) reported receipt of Fluarix Tetra (GSK), and 49 (11.1%) reported receipt of TetrAnflu (Sinovac). We matched 30–40 recipients of each vaccine (Fluarix Tetra = 30, Vaxigrip trivalent = 40, TetrAnflu = 40) by age and sex. Among included participants in each vaccine group, the average age ranged from 45.7 to 46.4 years, and between 26% and 40% were male (Table 1). Self-reported presence of any chronic conditions ranged from 22% to 40% of participants from each group. Pre-vaccination samples were collected between 109 and 124 days before vaccination, and post-vaccination samples were collected between 27 and 28 days after vaccination in each group. There were no statistically significant differences in these characteristics between participants in each vaccine group (Table 1).

**Table 1.** Characteristics of study participants by influenza vaccine received in 2025/26.

|  | **Vaxigrip**<br>**(Sanofi)** | **Fluarix Tetra**<br>**(GSK)** | **TetrAnflu**<br>**(Sinovac)** | **p-value\*** |
| --- | --- | --- | --- | --- |
| Vaccine valency | Trivalent | Quadrivalent | Quadrivalent |  |
| Number of participants | 40 | 30 | 40 |  |
| Age, mean (SD) | 45.7 (10.6) | 46.4 (11.1) | 45.9 (10.5) | 0.96 |
| Male (n, %) | 16 (40.0) | 8 (26.7) | 16 (40.0) | 0.43 |
| Presence of any chronic medical conditions (n, %) | 9 (22.5) | 9 (30.0) | 16 (40.0) | 0.24 |
| Received vaccination in prior season (2024/25) (n, %) | 39 (97.5) | 29 (96.7) | 39 (97.5) | 0.97 |
| Pre-vaccination sampling times in days before vaccination (median [range]) | 115.0 [1.0, 178.0] | 109.0 [3.0, 177.0] | 123.5 [1.0, 171.0] | 0.96 |
| Post-vaccination sampling times in days after vaccination (median [range]) | 27.0 [21.0, 39.0] | 27.5 [21.0, 39.0] | 28.0 [21.0, 34.0] | 0.73 |

\* p-values calculated from ANOVA tests against a null hypothesis of equal values across all three groups. P- values <0.05 would indicate that there are statistically significant differences between the three groups.

We measured the differences between pre-vaccination geometric mean HAI antibody titers (GMT, post-vaccination geometric mean HAI antibody titers (GMT, post-vaccination mean-fold rises in HAI antibody titers, proportion of participants who achieved seroconversion, and proportion of participants achieving post-vaccination HAI antibody titers ≥40, and proportion of participants who achieved a four-fold or greater rise in HAI antibody titers (Table 2). All metrics were of similar magnitude across groups for the vaccine antigens, with a statistically significant difference in responses to A(H1N1) between the three groups, but no statistically significant differences on subsequent pairwise comparisons between the three vaccines. Adjustment for age, sex, and the interval between pre-vaccination sampling and vaccination did not change these findings. Results were comparable in a sensitivity analysis limited to participants for whom the pre-sample was obtained within 90 days of vaccination (data not shown).

**Table 2.** Levels of hemagglutination inhibition antibody (HAI) titers before and after receipt of 2025/26 influenza vaccination with different brands of inactivated influenza vaccines.

| Vaccine strain | Antibody titers and ratios | Vaxigrip (Sanofi) | Fluarix Tetra (GSK) | TetrAnflu (Sinovac) | p-value<sup>2</sup> |
| --- | --- | --- | --- | --- | --- |
| A/Victoria/4897/2022 (H1N1) | Pre-vaccination GMT (95% CI) | 19.0 (12.8, 28.1) | 16.6 (10.6, 26.1) | 13.9 (9.36, 20.6) | 0.84 |
|  | Post-vaccination GMT (95% CI) | 28.3 (18.0, 44.6) | 26.4 (15.8, 44.2) | 31.9 (19.0, 53.7) | 0.37 |
|  | Mean-fold rise (95% CI) | 1.49 (1.14, 1.95) | 1.59 (1.05, 2.39) | 2.30 (1.53, 3.46) | 0.17 |
|  | Seroconversion<sup>1</sup> (% , 95% CI) | 7.5.0% (0.6%, 17%) | 17% (5.6%, 35%) | 32% (19%, 49%) | <0.01 |
|  | Proportion ≥40 (% , 95% CI) | 48% (32%, 64%) | 50% (31%, 69%) | 55% (38%, 71%) | 0.79 |
| A/Croatia/10136RV/2023 (H3N2) | Pre-vaccination GMT (95% CI) | 7.58 (5.96, 9.63) | 8.31 (6.28, 11.0) | 5.64 (4.90, 6.50) | 0.11 |
|  | Post-vaccination GMT (95% CI) | 9.01 (6.57, 12.4) | 10.7 (7.46, 15.4) | 6.71 (5.10, 8.83) | 0.82 |
|  | Mean-fold rise (95% CI) | 1.19 (0.98, 1.44) | 1.29 (1.01, 1.64) | 1.19 (0.97, 1.45) | 0.83 |
|  | Seroconversion (% , 95% CI) | 5% (0.61%, 17%) | 10% (2.1%, 27%) | 7.5% (1.6%, 20%) | 0.72 |
|  | Proportion ≥40 (% , 95% CI) | 10% (2.8%, 24%) | 27% (12%, 46%) | 7.5% (1.6%, 20%) | 0.06 |
| B/Austria/1359417/2021 (Victoria lineage) | Pre-vaccination GMT (95% CI) | 35.4 (22.0, 57.1) | 44.9 (24.3, 82.8) | 30.8 (18.8, 50.7) | 0.47 |
|  | Post-vaccination GMT (95% CI) | 77.3 (48.4, 123) | 101 (61.6, 165) | 78.6 (49.7, 124) | 0.78 |
|  | Mean-fold rise (95% CI) | 2.18 (0.17, 28.1.44, 3.31) | 2.24 (0.26, 19.1.49, 3.38) | 2.55 (1.77, 3.67) | 0.83 |
|  | Seroconversion (% , 95% CI) | 22% (11%, 38%) | 30% (15%, 49%) | 25% (13%, 41%) | 0.78 |
|  | Proportion ≥40 (% , 95% CI) | 80% (64%, 91%) | 83% (65%, 94%) | 82% (67%, 93%) | 0.93 |
| B/Phuket/3073/2013 (Yamagata lineage)<sup>3</sup> | Pre-vaccination GMT (95% CI) | 111 (79.1, 156) | 94.0 (63.8, 139) | 84.3 (57.2, 124) | 0.40 |
|  | Post-vaccination GMT (95% CI) | 142 (103, 195) | 130 (91.2, 185) | 121 (92.6, 159) | 0.30 |
|  | Mean-fold rise (95% CI) | 1.27 (1.10, 1.48) | 1.38 (1.09, 1.74) | 1.44 (1.09, 1.90) | 0.72 |
|  | Seroconversion (%; 95% CI) | 2.5% (0.06%, 13%, 15%) | 13% (3.8%, 31%) | 12% (4.2%, 27%) | 0.14 |
|  | Proportion ≥40 (%; 95% CI) | 98% (87%, 100%) | 97% (83%, 100%) | 95% (83%, 99%) | 0.83 |

Notes:
<sup>1</sup> Seroconversion is defined as a four-fold rise in hemagglutination inhibition antibody titer post-vaccination or a rise from a titer of <10 to 40 or above.
<sup>2</sup> \*p-values calculated from ANOVA tests against a null hypothesis of equal values across all three groups. P-values <0.05 would indicate statistically significant differences among the three groups.
Abbreviations: GMT = geometric mean titer
<sup>3</sup> The modest rise against B/Phuket (Yamagata) after Vaxigrip reflects cross-reactivity with the B/Victoria-lineage component present in the trivalent vaccine.

## DISCUSSION

Our findings of comparable HAI antibody responses induced by TetrAnflu compared to Fluarix Tetra and Vaxigrip are consistent with other published reports. A large multicenter phase 3 non-inferiority trial conducted in Chile and the Philippines showed that the TetrAnflu quadrivalent vaccine met non-inferiority criteria against Vaxigrip Tetra for all four strains in individuals aged ≥3 years, with point estimates favoring higher GMTs and seroconversion rates for several antigens [6]. A related analysis of the adult subset from the same trial further confirmed robust humoral responses alongside induction of virus-specific T-cell responses [8]. Together, these studies and our real-world data reinforce the observation that TetrAnflu elicits immune responses that are comparable to other available egg-based inactivated influenza vaccines.

Our study participants were drawn from a longitudinal cohort of healthcare workers, a group recommended for annual influenza vaccination because of occupational exposure risk and the potential to transmit infection to vulnerable patients [9,10]. Local influenza vaccination uptake among healthcare workers in Hong Kong remains moderate at approximately 40– 50% [11,12], underscoring the practical value of comparative data on available vaccines in this occupational risk group.

Several potential limitations of our study should be noted. Assignment to vaccine brand was not randomized and therefore, residual confounding cannot be fully excluded despite adjustment. Sample sizes per arm of 30-40 individuals also limit statistical power to detect smaller differences, but was limited by the available budget. As this study was not designed to capture pre-vaccination baseline serum samples on or within 7 days of influenza vaccination, we may not have captured asymptomatic or undetected influenza virus infections between the collection of pre-vaccination samples and post-vaccination samples. Finally, because sera were already banked from the longitudinal cohort, participants were not randomized to vaccine brand at the time of vaccination; residual confounding is therefore possible despite matching and covariate adjustment.

Only hemagglutination-inhibition (HAI) antibody responses were measured in this study. Virus neutralization, neuraminidase inhibition, and cellular immune responses were not assessed and could potentially differ between the three vaccines. Nevertheless, antibody titers measured by the HAI assay remain the established correlate of protection for inactivated influenza vaccines [13] and are therefore expected to reflect the relative strength of protection conferred by these products. Finally, our results are most generalizable to healthy, working-aged adults rather than children, older adults, or immunocompromised individuals.

In conclusion, TetrAnflu, Fluarix Tetra, and Vaxigrip induced comparable HAI antibody responses against the shared 2025/26 vaccine strains in this cohort of adult healthcare workers in Hong Kong. These findings, obtained under real-world conditions with matched groups and covariate adjustment, align with prior evidence of non-inferior immunogenicity for Sinovac’s quadrivalent inactivated vaccine relative to established products [6]. The results support the interchangeability of these licensed egg-based vaccines for seasonal influenza prevention among healthcare workers and may inform local procurement and vaccination policies.

## Data Availability

All data produced in the present study are available upon reasonable request to the authors

## Acknowledgments

The authors thank Julie Au for technical support.

## Data Availability

The data that support the findings of this study are available from the corresponding author upon reasonable request.

## CRediT author contributions

Wey Wen Lim: Conceptualization, Data curation, Formal analysis, Investigation,

Methodology, Funding acquisition, Writing – original draft, Writing – review & editing.

Lisa Touyon: Investigation, Resources, Writing – review & editing.

Loretta Mak: Data curation, Investigation, Project administration, Writing – review & editing.

Samuel M. S. Cheng: Investigation, Methodology, Resources, Writing – review & editing.

Yiu Chung Lau: Formal analysis, Writing – review & editing.

Dennis K. M. Ip: Conceptualization, Writing – review & editing.

Malik Peiris: Conceptualization, Funding acquisition, Supervision, Writing – review & editing.

Benjamin J. Cowling: Conceptualization, Funding acquisition, Methodology, Supervision,

Writing – review & editing.

Sook-San Wong: Conceptualization, Investigation, Methodology, Supervision, Funding acquisition, Writing – review & editing.

